# High occurrence of transportation and logistics occupations among vascular dementia patients: an observational study

**DOI:** 10.1101/19005512

**Authors:** A.C. van Loenhoud, C. de Boer, K. Wols, Y.A. Pijnenburg, A.W. Lemstra, F.H. Bouwman, N.D. Prins, P. Scheltens, R. Ossenkoppele, W.M. van der Flier

**Author notes:** corresponding author; address: room PK -1 Z 039, Vrije Universiteit Amsterdam, Amsterdam UMC, De Boelelaan 1118, 1081 HV, Amsterdam, The Netherlands. Co-author email addresses: C. de Boer, K. Wols, Y.A. Pijnenburg, A.W. Lemstra, F.H. Bouwman, N.D. Prins, P. Scheltens, R. Ossenkoppele, W.M. van der Flier.

## Abstract

**Background:** Growing evidence suggests a role of occupation in the emergence and manifestation of dementia. Occupations are often defined by complexity level, although working environments and activities differ in several other important ways. We aimed to capture the multi-faceted nature of occupation through its measurement as a qualitative (instead of a quantitative) variable and explored its relationship with different types of dementia.

**Methods:** We collected occupational information of 2,121 dementia patients with various suspected etiologies from the Amsterdam Dementia Cohort (age: 67±8, 57% male, MMSE: 21±5). Our final sample included individuals with Alzheimer’s disease (AD) dementia (n=1,467), frontotemporal dementia (n=281), vascular dementia (n=98), Lewy Body disease (n=174) and progressive supranuclear palsy/corticobasal degeneration (n=101). Within the AD group, we used neuropsychological data to further characterize patients by clinical phenotypes. All participants were categorized into one of 11 occupational classes, across which we evaluated the distribution of dementia (sub)types with Chi^2^ analyses. We gained further insight into occupation-dementia relationships through post-hoc logistic regressions that included various demographic and health characteristics as explanatory variables.

**Results:** There were significant differences in the distribution of dementia types across occupation groups (Chi^2^=85.87, p<.001). Vascular dementia was relatively common in the Transportation/Logistics sector, and higher vascular risk factors partly explained this relationship. Alzheimer’s disease occurred less in Transportation/Logistics and more in Health Care/Welfare occupations, which related to a higher/lower percentage of males. We found no relationships between occupational classes and clinical phenotypes of AD (Chi^2^=53.65, n.s.).

**Conclusions:** Relationships between occupation and dementia seem to exist beyond complexity level, which offers new opportunities for disease prevention and improvement of occupational health policy.

## Background

Dementia currently affects 46.8 million people worldwide [1], and this number is expected to increase as the aging population grows. Causes of dementia include Alzheimer’s disease (AD), frontotemporal dementia (FTD), vascular dementia (VaD) and movement disorders such as Lewy Body disease (DLB), progressive supranuclear palsy (PSP) and corticobasal degeneration (CBD). Apart from age and genetic influences, lifestyle factors play a role in the emergence and manifestation of dementia. For example, a comprehensive body of research points to the influence of education and occupation on dementia incidence [2–5]. Occupation could be an especially strong contributing factor, as most individuals spend a major part of their life at work.

Studies on the relationship between occupation and dementia have generally focused on the role of occupational complexity (e.g. rated as “white versus blue collar” [6]). Results indicate that older individuals who engaged in greater levels of occupational complexity have a better current cognitive status [6–13], a lower risk to develop dementia [5,14–17] or a different rate of clinical progression after the onset of dementia [4,11,18]. However, these studies mostly investigated one particular form of dementia, or “dementia” in general without specifying its type. Moreover, the common quantification of occupation by complexity level represents an oversimplification of the multi-faceted quality of this lifestyle factor. Working environments and activities differ in several important ways beyond complexity level, such as the level of physical activity, stress, social demands and exposure to hazardous substances [19,20]. Although a broader evaluation of occupation could thus provide important new insights in the context of dementia, this approach is relatively rare.

To overcome this scarcity in the literature, we took a different approach by i) measuring occupation as a qualitative (i.e. occupational class) rather than a quantitative variable (i.e. degree of occupational complexity), without making prior assumptions about complexity or other potential protective or harmful factors, and ii) exploring the relationships between occupation and various forms of dementia (i.e. different suspected etiologies and clinical phenotypes). Our sample included >2000 memory clinic patients with dementia due to AD, FTD, VaD or movement disorders. Occupation was categorized into several nominal groups (e.g. Pedagogical, Agricultural, Transportation/Logistics), across which we compared the distribution of these dementia types [21]. Within the AD subsample, we had sufficient data to additionally investigate relationships between occupation and specific cognitive profiles. As this study was exploratory in nature, we did not formulate a priori hypotheses. For relationships that emerged, we gained further insight through post-hoc analyses with additional demographic and health characteristics (i.e. age, sex, education and vascular risk factors [VRFs]).

## Methods

### Participants

Participants were selected from the Amsterdam Dementia Cohort (ADC [22,23]). The sample consists of individuals who visited the Alzheimer Center of the Amsterdam University Medical Center between 2000 and 2017. They underwent a standardized screening process that included the (patient and informant based) collection of demographic information, medical history, neurological examination, neuropsychological assessment, standard laboratory tests, brain magnetic resonance imaging (MRI) and lumbar puncture in a subsample. Diagnosis was established in a multidisciplinary meeting, based on common clinical criteria [24–34]. For some dementia types, diagnostic criteria underwent revisions during the period across which our participants were selected (Supplementary Table 1), but changes with respect to their core clinical characteristics were minor. Dementia diagnoses in the present study included AD, FTD, VaD, DLB and PSP/CBD.

### Assignment of occupational codes

Information on occupation was collected by a physician during a semi-structured interview with the participants and their informants. We subsequently coded this information according to the Dutch ROA-CBS 2014 occupational categorization system (BRC 2014), a derivative of the International Standard Classification of Occupations 2008 (ISCO 2008) [21]. The BRC 2014 consists of the following occupational classes: 1) Pedagogical, 2) Creative/Linguistic, 3) Commercial, 4) Business/Administrative, 5) Management, 6) Governmental/Law/Safety, 7) Technical, 8) Information and Communication Technology (ICT), 9) Agricultural, 10) Health Care/Welfare, 11) Service, 12) Transportation/Logistics (see Additional file 1). The BRC 2014 additionally includes a 13^th^ “Other” category (mostly applicable when occupational descriptions are absent or unclear), and makes further divisions into segments and subgroups. For the present paper, we were only interested in the occupational classes (i.e. classification at the segment level would result in 40 groups of insufficient size). We excluded participants in the “Other” category and those classified as “no occupation”. When multiple occupations were listed at once for a given participant (which is not permitted in our Chi^2^ model as described below in the “Statistical analyses” section [35]), we used the first job title for consistency reasons.

The assignment of BRC 2014 codes was performed by two raters (A.C.v.L and K.W.). To promote interrater consistency, a subset of the sample was independently coded and subsequently compared. Discordant cases were discussed with R.O. to reach consensus. The remaining sample was divided among A.C.v.L. and K.W., and only ambiguous cases were discussed and coded together. In rare occasions where this did not resolve classification uncertainty, medical records were inspected for more occupational details. If this did not provide useful information, the participant was coded as 13 (unclear), hence excluded for further analysis.

### Sample selection

We initially selected all ADC individuals with a dementia diagnosis at the baseline visit and occupational data available. Exclusion criteria were: 1) age below 40 years, 2) presence of autosomal dominant mutations for a neurodegenerative disease. The original selection consisted of 2,310 participants, of which we excluded 140 individuals with occupational descriptions that were either indicative of unemployment (∼70%) or non-informative (∼30%). Unemployed participants were beyond the scope of our study, as they constitute a heterogeneous group and our current interest was specifically in the distribution of dementia types among people who were part of the active workforce. Regarding non-informative descriptions, examples are “retired” (i.e. it gives no information about occupation beyond stating that the person is no longer employed) or “studied psychology” (i.e. it describes education rather than occupation). In addition, two participants were excluded because their occupational descriptions were not detailed enough to assign them to a category, even after inspection of their medical records. Finally, we decided to omit the category ICT due to the small sample size (n=12). ICT is a relatively “young” sector that did not have a prominent position in the labor market during the period in which the majority of our sample was employed. Similarly, 35 patients with relatively rare forms of dementia (e.g. alcohol-related dementias, normal pressure hydrocephalus, Creutzfeldt–Jakob disease) were excluded (Figure 1). The final sample included 2,121 individuals, with a diagnosis of AD (n=1,467), FTD (n=281), VaD (n=98), DLB (n=174) or PSP/CBD (n=101).

**Figure 1.**
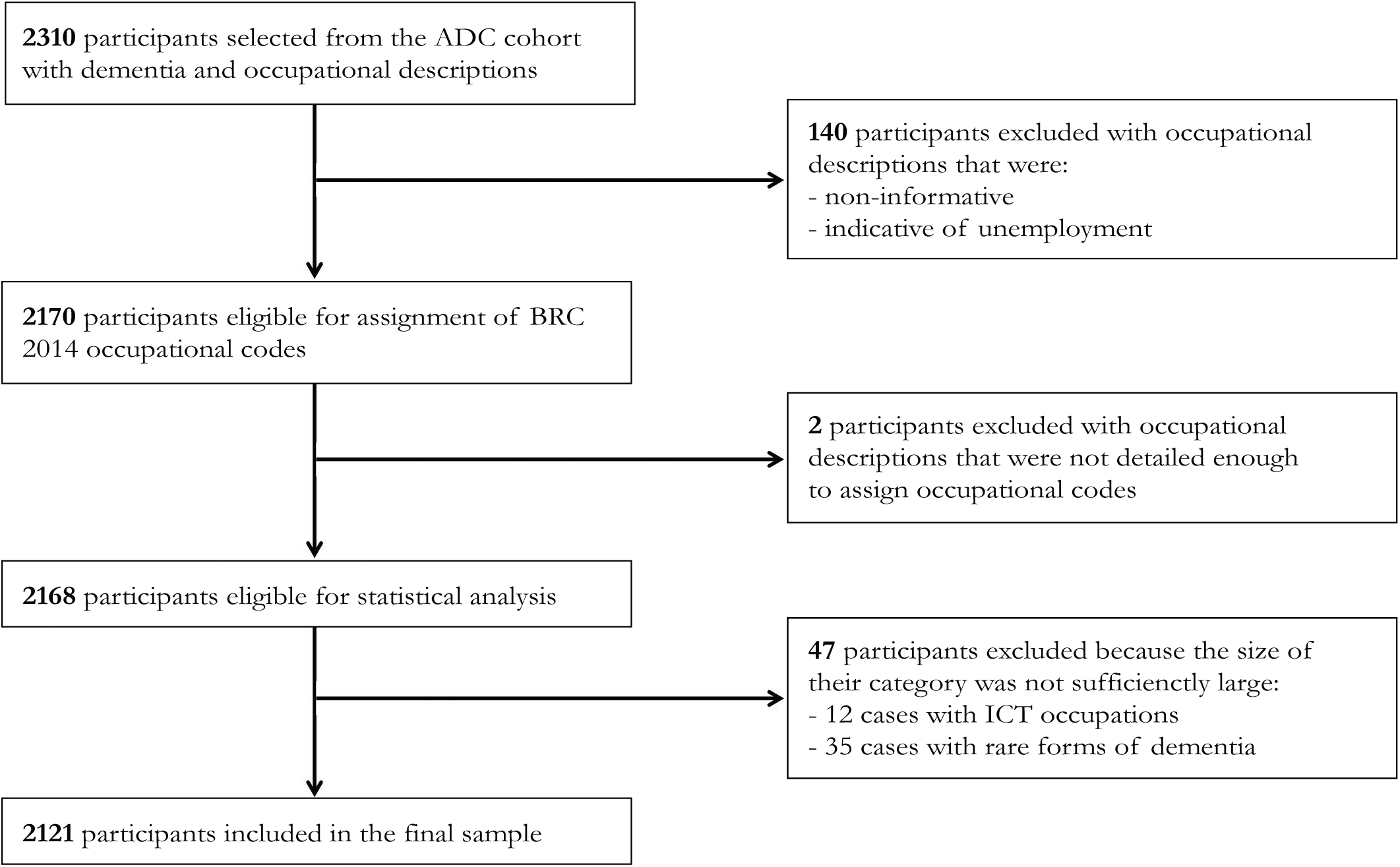
Flow diagram of the study sample. ADC=Amsterdam dementia cohort, BRC 2014=Dutch ROA-CBS 2014 occupational categorization system, a derivative of the Standard Classification of Occupations 2008 (ISCO 2008), ICT= Information and Communication Technology.

### Vascular risk factors

Complete data on VRFs were available for the majority of our sample (n=1,562/2,121), and included clinically measured body mass index (BMI), smoking, alcohol use (units per day), and presence or absence of a myocardial infarction, hypertension, hypercholesterolemia and diabetes mellitus in the medical history. We dichotomized BMI (≤25=0, >25=1), smoking (never smoked=0, ever smoked=1) and alcohol (0-1 units per day=0, >1 units per day=1) in order to derive a VRF score based on the sum of these seven risk factors [36].

### Neuropsychological assessment

Nearly all individuals underwent an elaborate neuropsychological assessment within three months of their diagnosis. For the AD dementia subsample, we therefore had a large dataset available (n=1,071/1,467) that consisted of participants who were cognitively tested in each of four cognitive domains: memory, attention/executive function, language and visuospatial function [37]. The <u>memory domain</u> included the total immediate and delayed recall as well as the recognition condition of the Rey Auditory Verbal Learning Test, and total recall in condition A of the Visual Association Test. In the <u>attention/executive function domain</u>, we used the Trail Making Test (part A and B), Stroop Test (condition I-III), 90-second Letter Digit Substitution Test [38], Digits Forward and Backwards, Letter Fluency and total score on the Frontal Assessment Battery. The <u>language domain</u> consisted of the Category Fluency, a short version of the Boston Naming Test and the total number of correctly named items of the Visual Association Test. Finally, the <u>visuospatial function domain</u> was assessed with the Dot Counting, Number Location and Fragmented Letters subtests of the Visual Object and Space Perception battery.

We carried out multiple imputation in SPSS 22.0 for Windows (SPSS, Chicago, IL, USA) to account for missing data (ranging from 1-45% per test). This procedure was performed using the fully conditional specification method, an iterative Markov Chain Monte Carlo approach suitable for arbitrary patterns of missing data. We imputed 25 datasets and included demographic, clinical, neuropsychological variables, APOE status and cerebrospinal fluid biomarkers as predictors in the model (i.e. occupational code was not used). In the pooled dataset, we created standardized residuals (i.e. W-scores) for each neuropsychological test score based on regressions with age, sex and educational level (measured on a seven-item scale, [39]). We calculated average W-scores across tests measuring memory, attention/executive function, language and visuospatial function, to obtain cognitive domain scores.

These cognitive domain scores were subsequently used to divide AD participants into clinical phenotypes, according to a previously validated method [40]. Specifically, we averaged across cognitive domains to create a “global cognition” composite score for each participant. Next, each cognitive domain score was dichotomized based on whether they were below a certain threshold relative to the global cognition score. A score of 1 thus means that a given cognitive domain “stands out” by being more affected than (the combination of) other domains. Based on these dichotomized scores, we assigned each participant to one of the following cognitive profiles: 1) memory-dominant, 2) attention/executive-dominant, 3) language-dominant, 4) visuospatial-dominant, 5) multi-domain (i.e. either more than one or no cognitive domain=1). As this profile categorization relies on which threshold is used to dichotomize the cognitive domain scores, we systematically tested all values between .250 and .500, and selected .255 as the “optimal” value that yielded 1) the lowest number of participants in the multi-domain profile, 2) the lowest sum of squared group sizes (Supplementary Figure 1). Most individuals had a memory-dominant cognitive profile (n=292), followed by a language-dominant (n=140), attention/executive-dominant (n=130) and visuospatial-dominant subgroup (n=126). The remaining participants fell into the multi-domain category (n=383).

### Statistical analyses

First, we carried out a Chi^2^ analysis in the total sample to investigate if the distribution of dementia types differed across occupational classes. In two sensitivity analyses, we reran the same model after exclusion of i) 93 persons with comorbid dementias (e.g. persons with AD as the primary diagnosis, but who also met NINDS-AIREN criteria for VaD) to create a sample with less overlap between diagnostic groups, and ii) 268 individuals who had reported more than one occupation, to enhance certainty that each participant was assigned to the most appropriate occupational class.

When Chi^2^ results were significant, we post-hoc inspected the adjusted residuals of each combination of occupation and dementia type. Adjusted residuals reflect the difference between the expected and observed count (e.g. the number of AD participants in the Pedagogical occupational class). An absolute value of ∼2 (i.e. 1.96 or −1.96) is considered significant at the p<.05 level. Since our contingency table was relatively large, we used a more stringent adjusted residual value threshold of ∼3 (i.e. 3.29 or −3.29 corresponds to p<.001; [41]). In addition, to assess whether the main relationships observed in significant Chi^2^ analyses could be explained by other demographic and health variables, we additionally performed logistic regressions. Specifically, we predicted dementia type from occupational class and evaluated age, sex, education and VRF score as potential explanatory variables. We used a forward selection procedure with a change-in-estimate (CIE) criterion of 10% [42,43] and ran these models in all participants with complete data (n=1,562/2,121).

Finally, we carried out Chi^2^ analyses to investigate whether occupation related to cognitive profiles within the AD subsample, following the same approach as described above. In a second analysis, we reran the same model using a different threshold for the dichotomization of cognitive domain scores (and subsequent categorization of participants into cognitive profiles; Supplementary Figure 1), and compared results to evaluate robustness of our findings.

## Results

### Participants

The mean age across diagnostic groups was 67±8 years (Table 1). Individuals with FTD were younger compared to other groups (63±7), and DLB participants were significantly older than the AD group (69±7). Overall, our sample included a somewhat larger proportion of males (57%), presumably reflecting the historically higher prevalence of males in the labor market (by comparison, the excluded “no occupation” sample was 82% female). The proportion of males was even higher among DLB participants (82%) in comparison with most other groups, while this was significantly lower for AD (52%) in contrast to all groups except PSP/CBD. Educational level was largely similar across dementia types in our sample (except for a DLB>VaD difference). Global cognitive impairment was most severe among AD participants (mean MMSE=20±5). Finally, VaD participants had a higher VRF score than most other diagnostic groups (p<.05; except FTD). Table 2 provides an overview of the number of participants in each occupational class for the total sample and according to dementia type. The largest occupation groups were Technical (n=429, 20%) and Business/Administrative (n=388, 18%), while the Agricultural class had the lowest number of participants (n=31, 1%).

**Table 1.** Participant characteristics in the total sample and according to dementia type

|  | **Total** | **AD** | **FTD** | **VaD** | **DLB** | **PSP/CBD** | **P-value** |
| --- | --- | --- | --- | --- | --- | --- | --- |
| **N** | 2,121 | 1,467 | 281 | 98 | 174 | 101 | - |
| **Age** | 67.20 ± 8.19 | 67.42 ± 8.35 | 63.92 ± 7.49 | 69.67 ± 8.02 | 69.37 ± 7.19 | 66.96 ± 6.75 | <.001 |
| **Sex (% male)** | 57.2 | 52.2 | 63.7 | 67.3 | 81.6 | 60.4 | <.001 |
| **Education (Verhage)<sup>a,b</sup>** | 5 (2) | 5 (2) | 5 (1) | 5 (1) | 6 (2) | 5 (2) | .007 |
| **MMSE<sup>b</sup>** | 21.25 ± 5.29 | 20.42 ± 5.14 | 23.48 ± 5.48 | 22.80 ± 4.78 | 22.42 ± 4.94 | 23.82 ± 4.82 | <.001 |
| **BMI<sup>c</sup>** | 25.24 ± 4.05 | 24.83 ± 3.95 | 26.88 ± 4.38 | 26.92 ± 4.70 | 25.10 ± 3.38 | 25.73 ± 3.58 | <.001 |
| **Smoking (% ever)<sup>c</sup>** | 53.3 | 52.0 | 56.5 | 55.2 | 54.2 | 59.6 | .464 |
| **Alcohol use (units/day)<sup>c</sup>** | .32 ± .47 | .32 ± .47 | .36 ± .48 | .22 ± .42 | .30 ± .46 | .28 ± .45 | .190 |
| **Myocard infarction (%)<sup>c</sup>** | 3.9 | 3.4 | 4.9 | 14.6 | 2.5 | 1.0 | <.001 |
| **Hypertension (%)<sup>c</sup>** | 26.3 | 25.3 | 23.2 | 54.6 | 22.0 | 28.6 | <.001 |
| **Hypercholesterolemia (%)<sup>c</sup>** | 9.3 | 9.3 | 8.2 | 9.4 | 10.4 | 9.2 | .957 |
| **Diabetes mellitus (%)<sup>c</sup>** | 9.5 | 8.0 | 11.5 | 29.2 | 10.9 | 5.1 | <.001 |
| **VRF score** | 1.97 ± 1.64 | 1.86 ± 1.62 | 2.00 ± 1.56 | 3.29 ± 1.82 | 2.03 ± 1.69 | 1.98 ± 1.46 | <.001 |

Data represent mean ± SD unless indicated otherwise. <sup>a</sup>Data are displayed as mode (interquartile range), <sup>b</sup>This information was available for 96% of the sample, <sup>c</sup>This information was available for 75% of the sample, <sup>d</sup>This information was available for 26% of the sample. AD=Alzheimer's disease dementia, FTD=frontotemporal dementia, VaD=vascular dementia, DLB=Lewy Body disease, PSP=progressive supranuclear palsy, CBD=corticobasal degeneration. MMSE=Mini-Mental State Examination, BMI=body mass index, VRF=vascular risk factors (total number).

**Table 2.** Distribution of dementia types across occupational classes.

|  | Total | AD | FTD | VaD | DLB | PSP/CBD |
| --- | --- | --- | --- | --- | --- | --- |
| **Total (n, [%])** | 2,121 (100) | 1,467 (69) | 281 (13) | 98 (5) | 174 (8) | 101 (5) |
| **Pedagogical (n, [%])** | 183 (100) | 129 (70) | 25 (14) | 7 (4) | 14 (8) | 8 (4) |
| **Creative/Linguistic (n, [%])** | 74 (100) | 53 (72) | 8 (11) | 4 (5) | 7 (9) | 2 (3) |
| **Commercial (n, [%])** | 205 (100) | 146 (71) | 29 (14) | 6 (3) | 17 (8) | 7 (3) |
| **Business/Administrative (n, [%])** | 388 (100) | 275 (71) | 48 (12) | 16 (4) | 28 (7) | 21 (5) |
| **Management (n, [%])** | 206 (100) | 145 (70) | 17 (8)* | 3 (1)* | 25 (12)* | 16 (8)* |
| **Governmental/Law/Safety (n, [%])** | 90 (100) | 64 (71) | 10 (11) | 2 (2) | 9 (10) | 5 (6) |
| **Technical (n, [%])** | 429 (100) | 273 (64)* | 72 (17)* | 28 (7)* | 39 (9) | 17 (4) |
| **Agricultural (n, [%])** | 31 (100) | 16 (52)* | 7 (23) | 1 (3) | 3 (10) | 4 (13)* |
| **Health Care/Welfare (n, [%])** | 283 (100) | 222 (78)*** | 26 (9)* | 8 (3) | 13 (5)* | 14 (5) |
| **Service (n, [%])** | 141 (100) | 97 (69) | 23 (16) | 11 (8) | 7 (5) | 3 (2) |
| **Transportation/Logistics (n, [%])** | 91 (100) | 47 (52)*** | 16 (18) | 12 (13)*** | 12 (13) | 4 (4) |

10 cells (18.2%) had an expected count less than 5; the minimum expected count was 1.43. AD=Alzheimer's disease dementia, FTD=frontotemporal dementia, VaD=vascular dementia, DLB=Lewy Body disease, PSP=progressive supranuclear palsy, CBD=corticobasal degeneration. \*Chi<sup>2</sup> adjusted residual ≤ -2 or ≥2 (corresponding to p<.05), \*\*\*Chi<sup>2</sup> adjusted residual ≤ -3 or ≥3 (corresponding to p<.001).

### Relationship between occupation and dementia types

There were significant differences in the distribution of dementia types across occupation groups (Chi^2^=85.87, p<.001, Table 2). The adjusted residuals revealed three effects significant at p<.001. First, individuals from the Transportation/Logistics sector were more often diagnosed with VaD (adjusted residual: 4.0). Second, this occupational class had fewer AD participants (adjusted residual: −3.7). Third, in the Health Care/Welfare group, AD was relatively common (adjusted residual: 3.6). These results are displayed in Figure 2, along with 11 other effects (e.g. a high proportion of PSP/CBD cases in the Agricultural sector) that were significant at p<.05 (adjusted residuals >2 or <-2). When we repeated the Chi^2^ analysis with exclusion of 93 individuals with comorbid dementias (n=2,028), results remained essentially unchanged (Chi^2^=78.73, p<.001; Supplementary Table 2). The same was true for results based on a sample that excluded 268 individuals with multiple occupations (n=1,853; Chi^2^=79.93, p<.001; Supplementary Table 3).

**Figure 2.**
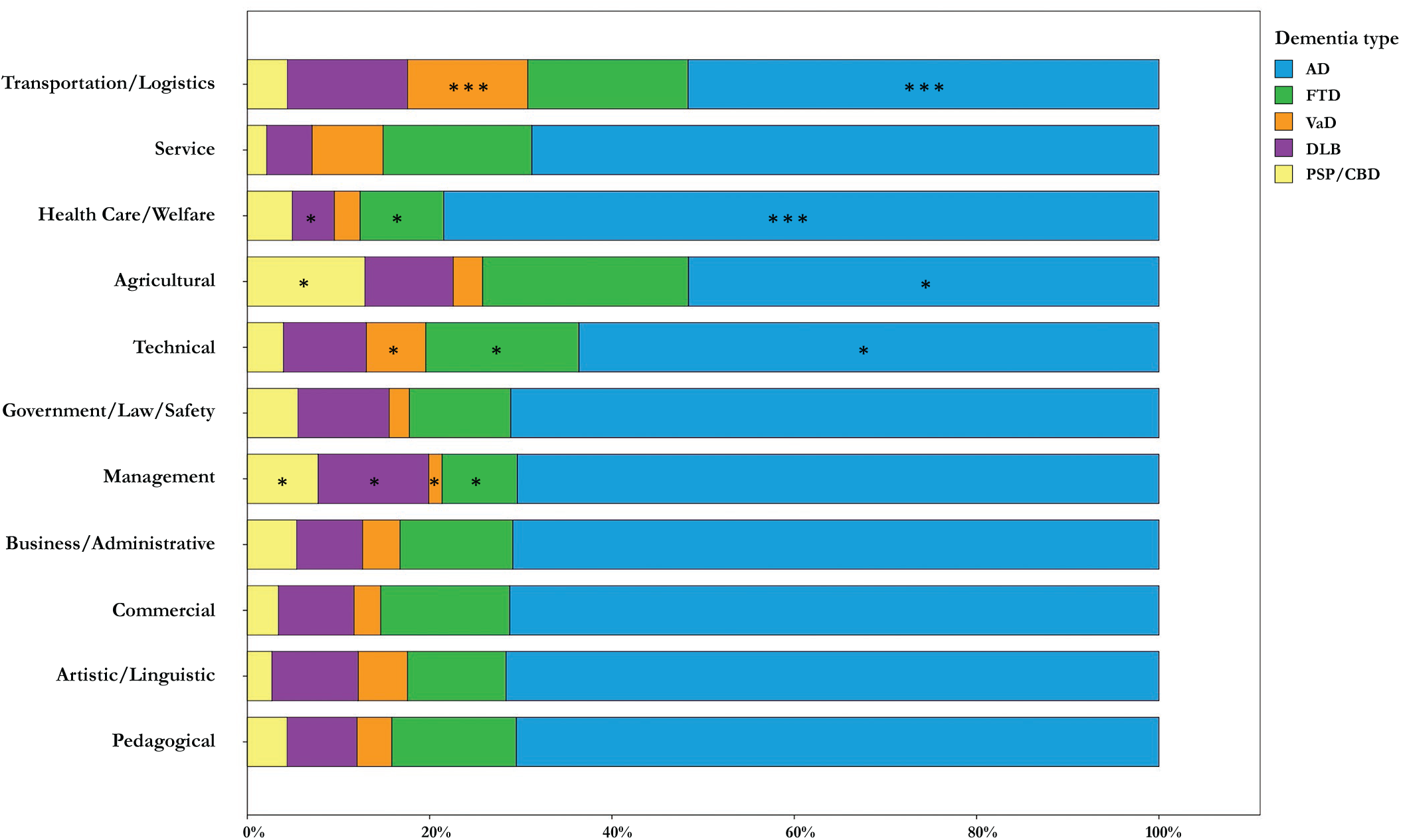
Proportions of dementia types for each occupational class. AD=Alzheimer’s disease dementia, FTD=frontotemporal dementia, VaD=vascular dementia, DLB=Lewy Body disease, PSP=progressive supranuclear palsy, CBD=corticobasal degeneration. *Chi^2^ adjusted residual ≤ −2 or ≥2 (corresponding to p<.05), ***Chi^2^ adjusted residual ≤ −3 or ≥3 (corresponding to p<.001).

### Contribution of age, sex, education and vascular risk factors

In line with the previous results, uncorrected logistic regression models with either AD or VaD as a dependent variable (AD/VaD=1 in separate models, other diagnosis=0) confirmed Transportation/Logistics to be significantly related to both VaD (β=1.23, odds ratio [OR] = 3.41, p<.01) and AD (β=-.85, OR=.43, p<.001). Likewise, Health Care/Welfare was a significant determinant of AD (β=.55, OR=1.74, p<.01; Supplementary Table 4). Forward selection of explanatory variables in corrected models revealed that VRF score, but no other factors (i.e. age, sex, education), was positively associated with a VaD diagnosis (β=.43, OR=1.53, p<.001) and reduced the Transportation/Logistics effect by 15% (from β=1.23 to 1.04, OR=3.41 to 2.84). For AD, only the variable sex (range β=.68-.69, OR=1.96-1.98, p<.001) significantly reduced the effects of Transportation/Logistics (from β=-.85 to −.62, OR=.43 to .54; 27%) and Health Care/Welfare (from β=.55 to .31, OR=1.74 to 1.37; 43%). Figure 3 gives an overview of the number of VRFs and percentage of males for all dementia types and occupational classes.

**Figure 3.**
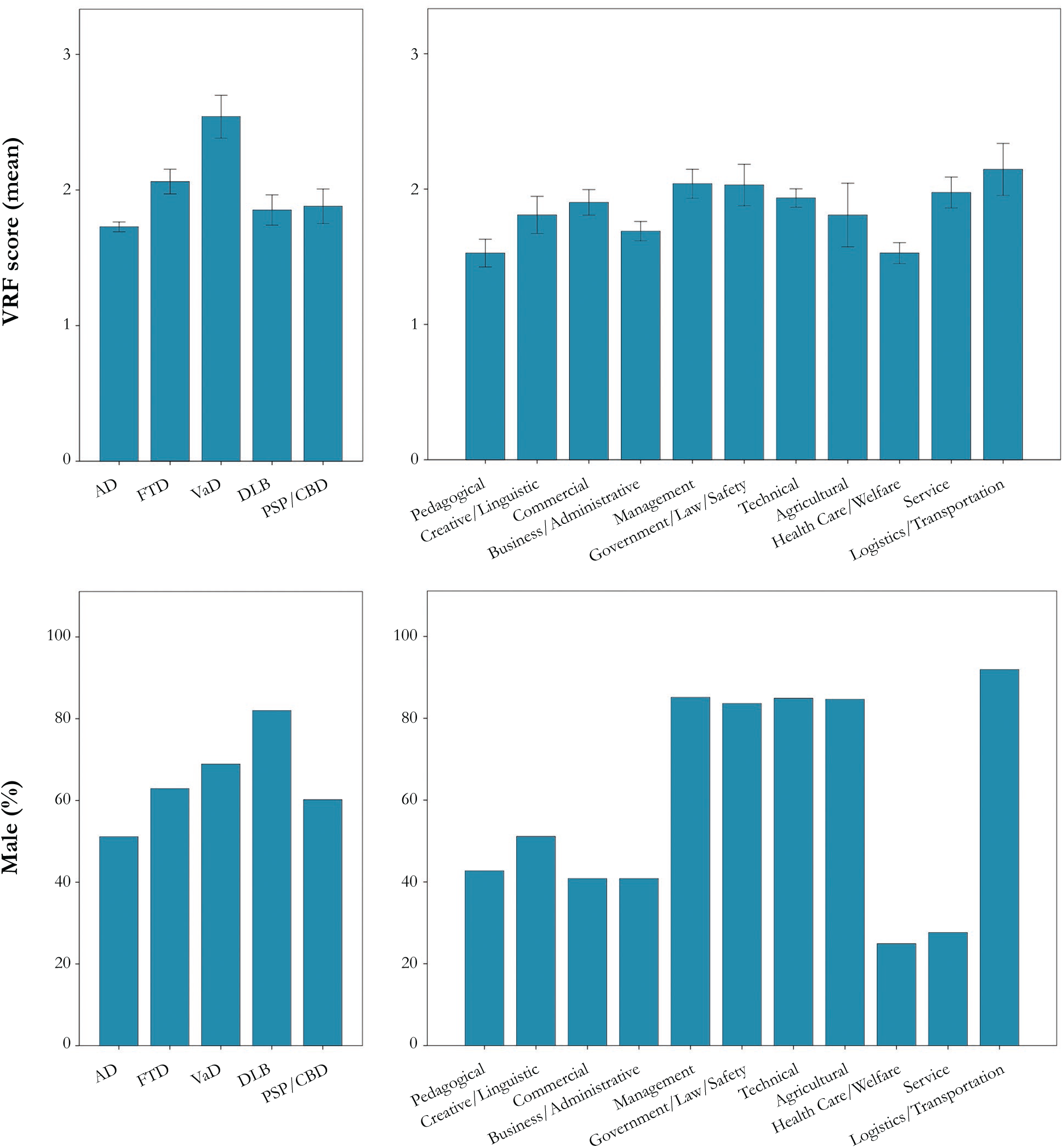
Mean number of vascular risk factors and male percentage for occupational classes and dementia types. Error bars represent standard errors. AD=Alzheimer’s disease dementia, FTD=frontotemporal dementia, VaD=vascular dementia, DLB=Lewy Body disease, PSP=progressive supranuclear palsy, CBD=corticobasal degeneration. VRF=vascular risk factors, including included body mass index (BMI), smoking, alcohol use (units per day), and presence or absence of a myocardial infarction, hypertension, hypercholesterolemia and diabetes mellitus in the medical history.

### Relationship between occupation and cognitive profiles of AD

We found only trend-significant differences between occupations in the distribution of cognitive profiles (Chi^2^=53.65, p=.07; Table 3). Supplementary Figure 2 provides an overview of these findings. In a secondary analysis, we repeated the same analysis using cognitive profiles that were calculated based on a different threshold of −.442 (Supplementary Figure 1). Similar to the original findings, these results were not significant (Chi^2^=38.24, p=.55).

**Table 3.**
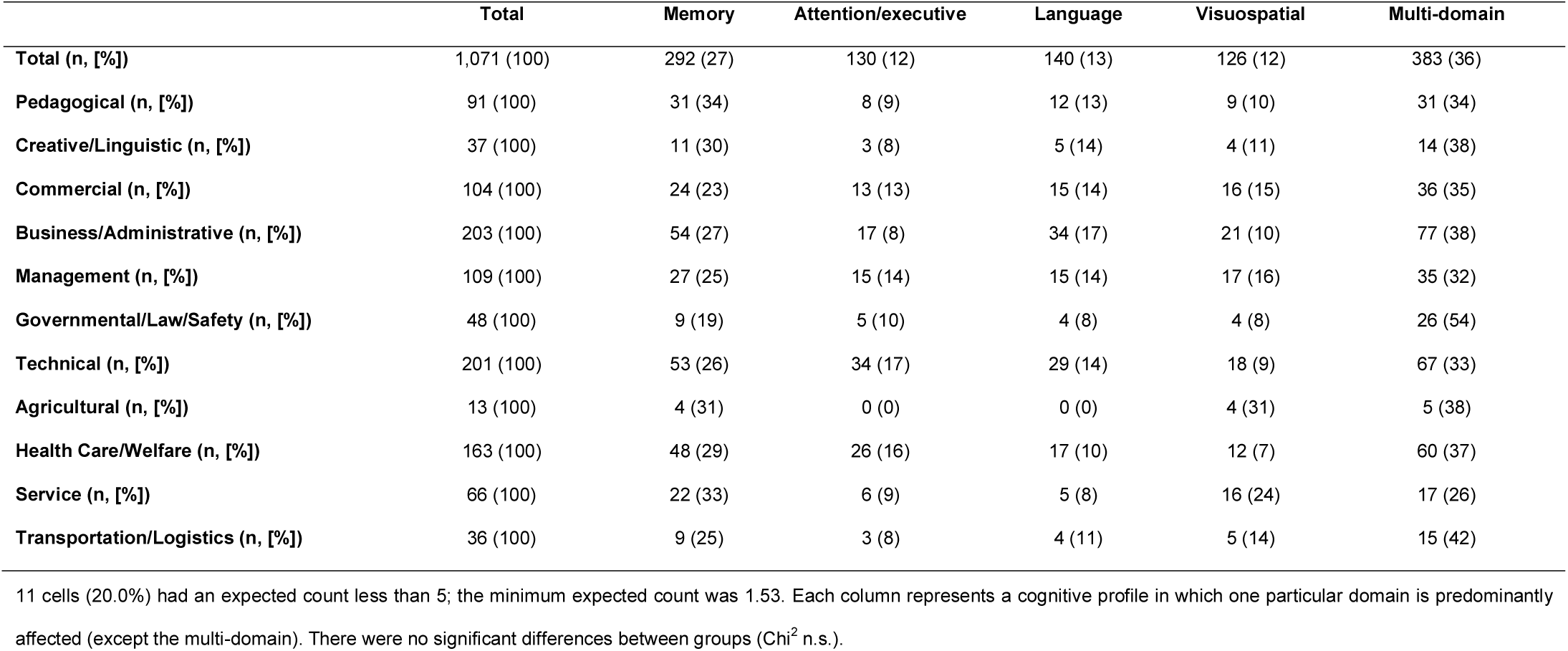
Distribution of cognitive profiles of AD across occupational classes.

## Discussion

In this exploratory study, we investigated the association between occupation and dementia (sub)types in a large sample of memory-clinic patients. The main finding was that VaD was relatively common in participants with Transportation/Logistics occupations. Post-hoc analyses suggested that this observed relationship was partly explained by a higher number of VRFs among these individuals. In addition, the number of AD participants was relatively low in the Transportation/Logistics sector, and high among Health Care/Welfare occupations. We found that these effects were mostly attributable to differences between groups in sex distribution. There was no relationship between occupation types and AD-related cognitive profiles.

Our finding that VRFs are more prominent among VaD participants is in line with previous studies [44,45], but of particular interest was our observation of a higher number of VRFs in Transportation/Logistics occupations. Although we cannot make any inferences about dementia risk (due to the absence of a healthy control group), it raises the possibility that certain job-related aspects or occupational habits contribute to the development of VRFs and the emergence of VaD. Our data at least warrant an increased awareness and monitoring of VRFs by occupational physicians in the Transport/Logistics sector. Furthermore, the lower presence of AD cases in Transpiration/Logistics groups (male-predominant) versus higher AD occurrence Healthcare/Welfare (female-predominant), is in line with evidence that women have a higher risk for AD than men [46,47]. Importantly, while VRFs and sex partially explained the main occupation-dementia relationships we observed, these factors did not account for all variance. This suggests there might be other important ways in which occupations are related to the emergence and manifestation of dementia.

In contrast to our present study, most previous studies operationalized occupation by complexity level. Apart from coarse ratings such as “white versus blue collar”, more fine-grained classifications have been made in which the complexity of multiple occupational aspects was considered. An example is the use of questionnaires that differentiated between working with “people, data or things” (e.g. [12]). However, although these approaches have provided important insights, they did not consider other potential job-related risks or protective factors. The few studies that did take a more qualitative approach towards the measurement of occupation have used “job exposure matrices” or “occupational description databases” to show, among other results, that dementia risk and late-life cognition were influenced by the degree of work control (e.g. influence over work planning, variation in task content) [48], level of human interaction and physical activity [49] and mental demands (e.g. information processing, pattern detection, creativity) [50].

The novelty of the present work lies our “assumption-free” approach that did not involve any additional characterization of occupations besides their categorization across 11 occupational classes. This enabled examining relationships between occupation and dementia (sub)types in an exploratory manner. Importantly, occupational classes and complexity level did not correspond in a straightforward manner. Most classes comprised occupations that varied considerably with respect to complexity (e.g. the “Technical” group both contained engineers and assembly workers, see Additional file 1). Another strength of our study is the fact that our sample consisted of dementias with different suspected etiologies. Most earlier studies have either focused on the occurrence of dementia as a general syndrome or one particular form of dementia. As our dataset included over two thousand carefully diagnosed participants, we were able to create several independent diagnostic groups and AD-related cognitive profiles that were sufficiently sized for statistical analysis. Finally, we consider use of the BRC 2014 classification system to be an important advantage, because it improves the replicability of our findings across other populations on an international scale.

Our study has several limitations. First, although we collected occupational data with a refined method that involved a semi-structured interview with the patient and caregiver, we did not have detailed enough information to capture a person’s entire occupational timeline. For individuals who had multiple occupations listed, we often could not retrieve their sequence and respective durations. It is therefore possible that the first-mentioned job not always reflect the person’s foremost/primary occupation, but rather the most recent occupation. This is non-optimal because of potential reverse causation: we cannot rule out that some patients switched jobs as a consequence of their dementia diagnosis (e.g. choosing less demanding work in a different sector). However, we have no reason to assume that reverse causation is more prominent for some dementia (sub)types than others, and thus the degree of bias seems limited. Moreover, our results did not change after exclusion of individuals with multiple occupational descriptions. A second limitation concerns our inability to define occupation groups beyond the highest hierarchical level of the BRC 2014, due to a lack of detail in occupational descriptions and the fact that the resulting number of categories would be too large for our sample size (i.e. ∼40 occupation groups). More detailed distinctions between occupations would have decreased heterogeneity within categories, and might have revealed additional relevant occupation-dementia relationships that were currently missed. Third, the absence of a healthy control group restricted our ability to make inferences about dementia risk or distinguish between protective and harmful effects of occupation (Supplementary Figure 3), which limits the interpretability of our results. Fourth, our sample of dementia patients may not be fully representative of the population of individuals with dementia as a whole. As not every person with dementia is referred to a memory clinic or seeks medical care in an academic expertise center such as the Alzheimer Center Amsterdam, the generalizability of our results is somewhat limited. Fifth, although diagnoses of dementia were generally based on both clinical observation and in vivo neuroimaging biomarkers, we did not have neuropathological confirmation of the underlying etiologies. It is therefore possible that some participants were categorized under a dementia type that would not completely correspond with their neuropathological diagnosis. Finally, although our total sample was large (N=2,121), some occupational classes (e.g. Agricultural) contained a low number of observations. It is possible that we have failed to find certain relationships with dementia types due to limited power in these classes.

## Conclusions

To conclude, we observed relationships between occupation and dementia types. Our findings suggest that these relationships emerged beyond occupational complexity level. Differences between occupational classes in the number of VRFs and sex distributions partially explained our results, but other - currently unknown - factors likely play a role as well. It is important to note that associations between occupation and dementia could exist for multiple reasons. Whilst it seems plausible that some occupational characteristics causally relate to the occurrence of specific dementia types, another possibility is that certain (genetic or early environmental) factors both influence career choice and dementia development later in life. This is consistent with theoretical frameworks that propose a neurodevelopmental component in the etiology of dementia [51–53]. Ultimately, as most people spend the majority of their life working, occupation could be an important lifestyle factor to consider in relation to preventive strategies for dementia. A better understanding of occupation-dementia relationships may improve occupational health policy through reduction of job hazards and more targeted health monitoring by occupational physicians.

## Data Availability

The datasets used and/or analyzed during the current study are available from the corresponding author on reasonable request.

## List of abbreviations

AD: Alzheimer’s disease
VaD: vascular dementia
FTD: frontotemporal dementia
DLB: Lewy Body disease
PSP: progressive supranuclear palsy
CBD: corticobasal degeneration
ADC: Amsterdam Dementia Cohort
MRI: magnetic resonance imaging
BRC 2014: Dutch ROA-CBS 2014 occupational categorization system
ISCO 2008: the International Standard Classification of Occupations 2008
ICT: Information and Communication Technology
BMI: body mass index
VRF: vascular risk factor
CIE: change-in-estimate
OR: odds ratio

## Declarations

### Ethics approval and consent to participate

Participants gave written informed consent to use their medical data for scientific purposes. This procedure was approved by the local Medical Ethics Committee.

### Consent for publication

Not applicable

### Authors’ contributions

A.C.v.L. was involved in the methodological design of the research project and assignment of occupational codes; she furthermore conducted the statistical analyses, interpreted the data and wrote the manuscript. C.d.B. was responsible for the selection of the participant data from the ADC. K.W. took part in the methodological design and the assignment of occupational codes. Y.A.P., A.W.L., F.H.B., N.D.P., and P.S. were involved in the standardized screening process of participants and collection of occupational data. R.O. was involved in the methodological design of the research project, interpretation of the data and writing the manuscript. W.M.v.d.F. is the coordinator of the ADC, and took part in the interpretation of the data and editing of the manuscript. All authors read and approved the final manuscript.

## Acknowledgements

Not applicable

---

**Supplementary Table 1.** Clinical criteria for different dementia types between 2000-2017.

|  |  |
| --- | --- |
| **AD** | McKhann et al. (NINCDS-ADRDA) [24]; McKhann et al. (NIA-AA) [25] |
| **FTD** | Neary et al. [26]; Rascovsky et al. (behavioral variant) [27] and Gorno-Tempini et al. (primary progressive aphasia) [28] |
| **VaD** | Román et al. (NINDS-AIREN) [29] |
| **DLB** | McKeith et al. [30]; McKeith et al [31] |
| **PSP** | Litvan et al. (NINDS-SPSP) [32] |
| **CBD** | Boeve et al. [33]; Armstrong et al. [34] |

For each dementia type, the clinical criteria used are provided in chronological order. AD=Alzheimer's disease dementia, FTD=frontotemporal dementia, VaD=vascular dementia, DLB=Lewy Body disease, PSP=progressive supranuclear palsy, CBD=corticobasal degeneration.

**Supplementary Table 2.** Distribution of dementia types across occupations groups after excluding comorbid cases.

|  | **Total** | **AD** | **FTD** | **VaD** | **DLB** | **PSP/CBD** |
| --- | --- | --- | --- | --- | --- | --- |
| **Total (n, [%])** | 2,028 (100) | 1,401 (69) | 274 (14) | 83 (4) | 171 (8) | 99 (5) |
| **Pedagogical (n, [%])** | 172 (100) | 122 (71) | 24 (14) | 5 (3) | 14 (8) | 7 (4) |
| **Creative/Linguistic (n, [%])** | 72 (100) | 52 (72) | 8 (11) | 4 (6) | 6 (8) | 2 (3) |
| **Commercial (n, [%])** | 195 (100) | 138 (71) | 28 (14) | 5 (3) | 17 (9) | 7 (4) |
| **Business/Administrative (n, [%])** | 374 (100) | 265 (71) | 46 (12) | 15 (4) | 27 (7) | 21 (6) |
| **Management (n, [%])** | 195 (100) | 135 (69) | 17 (9) | 3 (2) | 24 (12)* | 16 (8)* |
| **Governmental/Law/Safety (n, [%])** | 88 (100) | 63 (72) | 10 (11) | 1 (1) | 9 (10) | 5 (6) |
| **Technical (n, [%])** | 409 (100) | 261 (64)* | 69 (17)* | 24 (6)* | 39 (10) | 16 (4) |
| **Agricultural (n, [%])** | 30 (100) | 15 (50)* | 7 (23) | 1 (3) | 3 (10) | 4 (13)* |
| **Health Care/Welfare (n, [%])** | 271 (100) | 211 (78)*** | 26 (10)* | 7 (3) | 13 (5)* | 14 (5) |
| **Service (n, [%])** | 135 (100) | 94 (70) | 23 (17) | 8 (6) | 7 (5) | 3 (2) |
| **Transportation/Logistics (n, [%])** | 87 (100) | 45 (52)*** | 16 (18) | 10 (11)*** | 12 (14) | 4 (5) |

10 cells (18.2%) had an expected count less than 5; the minimum expected count was 1.23. AD=Alzheimer's disease dementia, FTD=frontotemporal dementia, VaD=vascular dementia, DLB=Lewy Body disease, PSP=progressive supranuclear palsy, CBD=corticobasal degeneration. \*Chi<sup>2</sup> adjusted residual is $\leq -2$ or $\geq 2$ (corresponding to $p < .05$ ), \*\*\*Chi<sup>2</sup> adjusted residual is $\leq -3$ or $\geq 3$ (corresponding to $p < .001$ ).

**Supplementary Table 3.** Distribution of dementia types across occupation groups after excluding cases with multiple occupations.

|  | Total | AD | FTD | VaD | DLB | PSP/CBD |
| --- | --- | --- | --- | --- | --- | --- |
| **Total (n, [%])** | 1,853 (100) | 1,282 (69) | 246 (13) | 83 (5) | 154 (8) | 88 (5) |
| **Pedagogical (n, [%])** | 155 (100) | 110 (71) | 21 (14) | 5 (3) | 11 (7) | 8 (5) |
| **Creative/Linguistic (n, [%])** | 62 (100) | 45 (73) | 8 (13) | 2 (3) | 5 (8) | 2 (3) |
| **Commercial (n, [%])** | 73 (100) | 120 (69) | 27 (16) | 5 (3) | 14 (8) | 7 (4) |
| **Business/Administrative (n, [%])** | 356 (100) | 250 (70) | 43 (12) | 15 (4) | 28 (8) | 20 (6) |
| **Management (n, [%])** | 199 (100) | 139 (70) | 17 (9)* | 3 (2)* | 24 (12)* | 16 (8)* |
| **Governmental/Law/Safety (n, [%])** | 83 (100) | 59 (71) | 9 (11) | 2 (2) | 9 (11) | 4 (5) |
| **Technical (n, [%])** | 360 (100) | 233 (65)* | 60 (17)* | 25 (7)* | 30 (8) | 12 (3) |
| **Agricultural (n, [%])** | 25 (100) | 12 (48)* | 6 (24) | 1 (4) | 3 (12) | 3 (12) |
| **Health Care/Welfare (n, [%])** | 245 (100) | 193 (79)*** | 23 (9) | 6 (2) | 12 (5)* | 11 (5) |
| **Service (n, [%])** | 117 (100) | 81 (69) | 19 (16) | 8 (7) | 7 (6) | 2 (2) |
| **Transportation/Logistics (n, [%])** | 78 (100) | 40 (51)*** | 13 (17) | 11 (14)*** | 11 (14) | 3 (4) |

10 cells (18.2%) had an expected count less than 5; the minimum expected count was 1.12. AD=Alzheimer's disease dementia, FTD=frontotemporal dementia, VaD=vascular dementia, DLB=Lewy Body disease, PSP=progressive supranuclear palsy, CBD=corticobasal degeneration. \*Chi<sup>2</sup> adjusted residual ≤ -2 or ≥2 (corresponding to p<.05), \*\*\*Chi<sup>2</sup> adjusted residual ≤ -3 or ≥3 (corresponding to p<.001).

**Supplementary Table 4.** Uncorrected and corrected logistic regression models of relationships between occupational class and dementia type.

|  | Uncorrected |  |  | Corrected |  |  |  |
| --- | --- | --- | --- | --- | --- | --- | --- |
| | $\beta$ | OR | P-value | $\beta$ | OR | P-value | CIE ($\beta$) |
| **VaD** |  |  |  |  |  |  |  |
| **Transportation/Logistics<sup>a</sup>** | 1.23 | 3.41 | .004 | 1.04 | 2.84 | .018 | 14.8 % |
| **AD** |  |  |  |  |  |  |  |
| **Transportation/Logistics<sup>b</sup>** | -.85 | .43 | .001 | -.62 | .54 | .020 | 27.4 % |
| **Health Care/Welfare<sup>b</sup>** | .55 | 1.74 | .003 | .31 | 1.37 | .101 | 43.2 % |

VaD=vascular dementia, AD=Alzheimer's disease dementia, OR=odds ratio, CIE=change-in-estimate. We used a forward selection procedure with a change-in-estimate (CIE) criterion of 10% [42,43] to select relevant covariates among all participants with complete data (n=1,562/2,121). We tested age, sex, education and VRFs as covariates; only VRFs and sex were finally included.

**Supplementary Figure 1.**
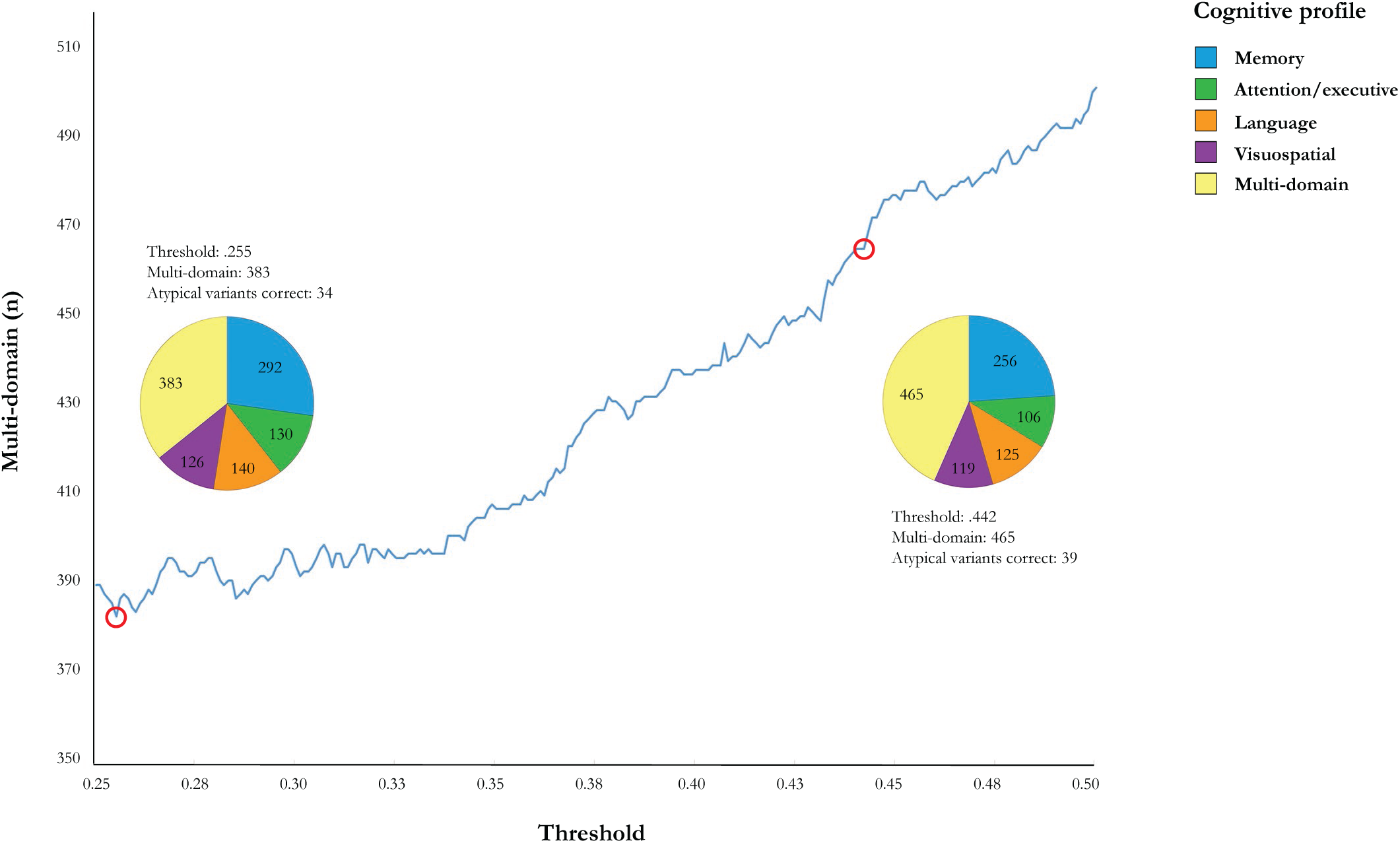
Thresholds used for the dichotomization of cognitive domains and division of participants across cognitive profiles. Within the AD subsample (n=1,071), we dichotomized each cognitive domain score (i.e. memory, attention/executive functions, language and visuospatial functions), based on whether or not a domain’s W-score was considerably lower compared to an individual’s global cognition score. We used an optimal threshold between .250 and .500 for this dichotomization, and defined “optimality” in two different ways: 1) the lowest number of participants in the multi-domain profile and the lowest sum of squared group sizes (threshold=.255), 2) the highest number of atypical variant AD cases (n=85) categorized into the language (i.e. for logopenic aphasia) or visuospatial cognitive profile (i.e. for PCA; n=.442). Note that neither thresholds (i.e. .255 or .442) resulted in all atypical variants being categorized into these two cognitive profiles, presumably because in advanced disease stages, several cognitive domains become affected. For many participants with an initial logopenic aphasia or PCA diagnosis, visuospatial/language may no longer have been predominant at study inclusion, causing them to be assigned to the multi-domain instead.

**Supplementary Figure 2.**
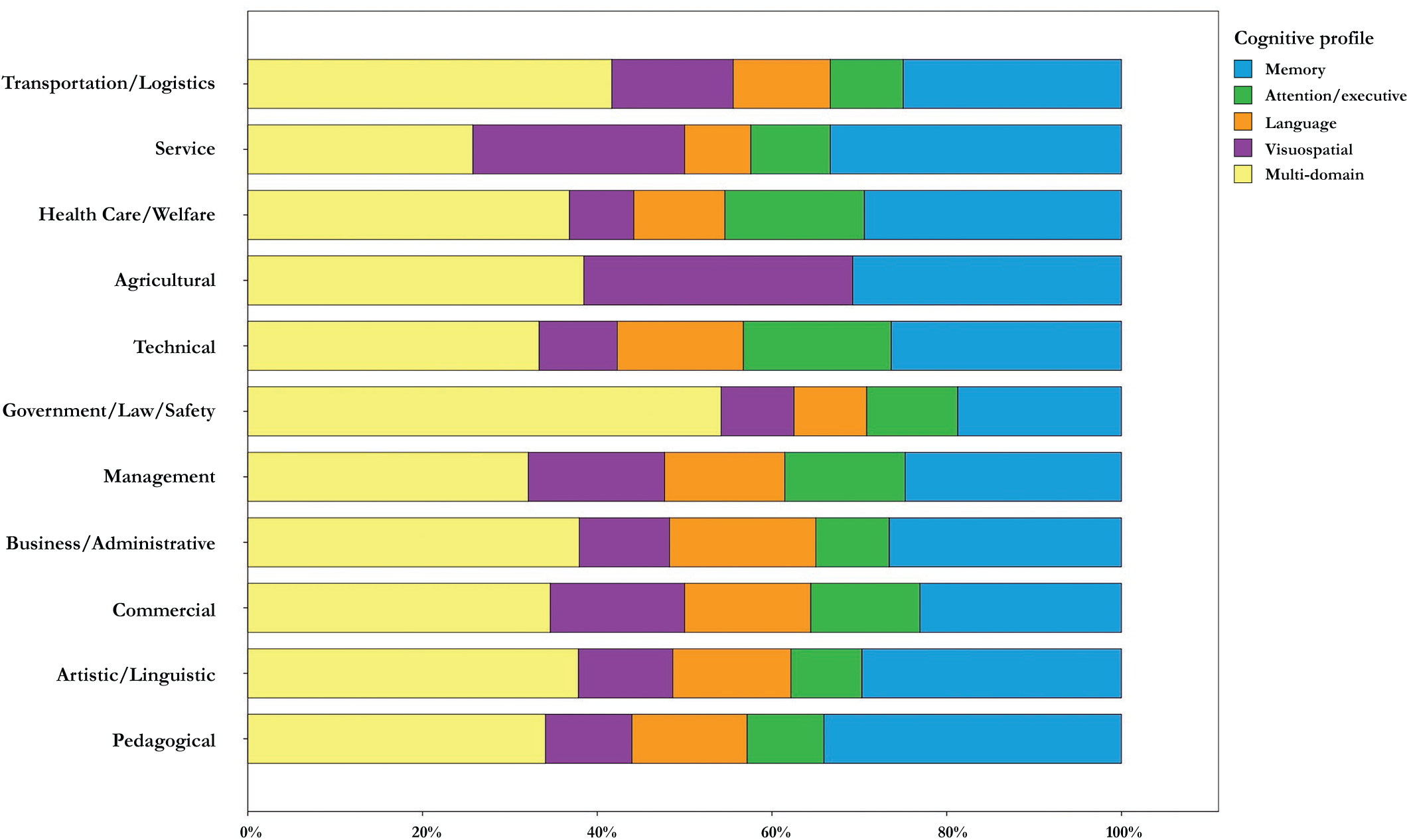
Proportions of AD-related cognitive profiles for each occupational class. There were no significant differences between groups (Chi^2^ n.s.).

**Supplementary Figure 3.**
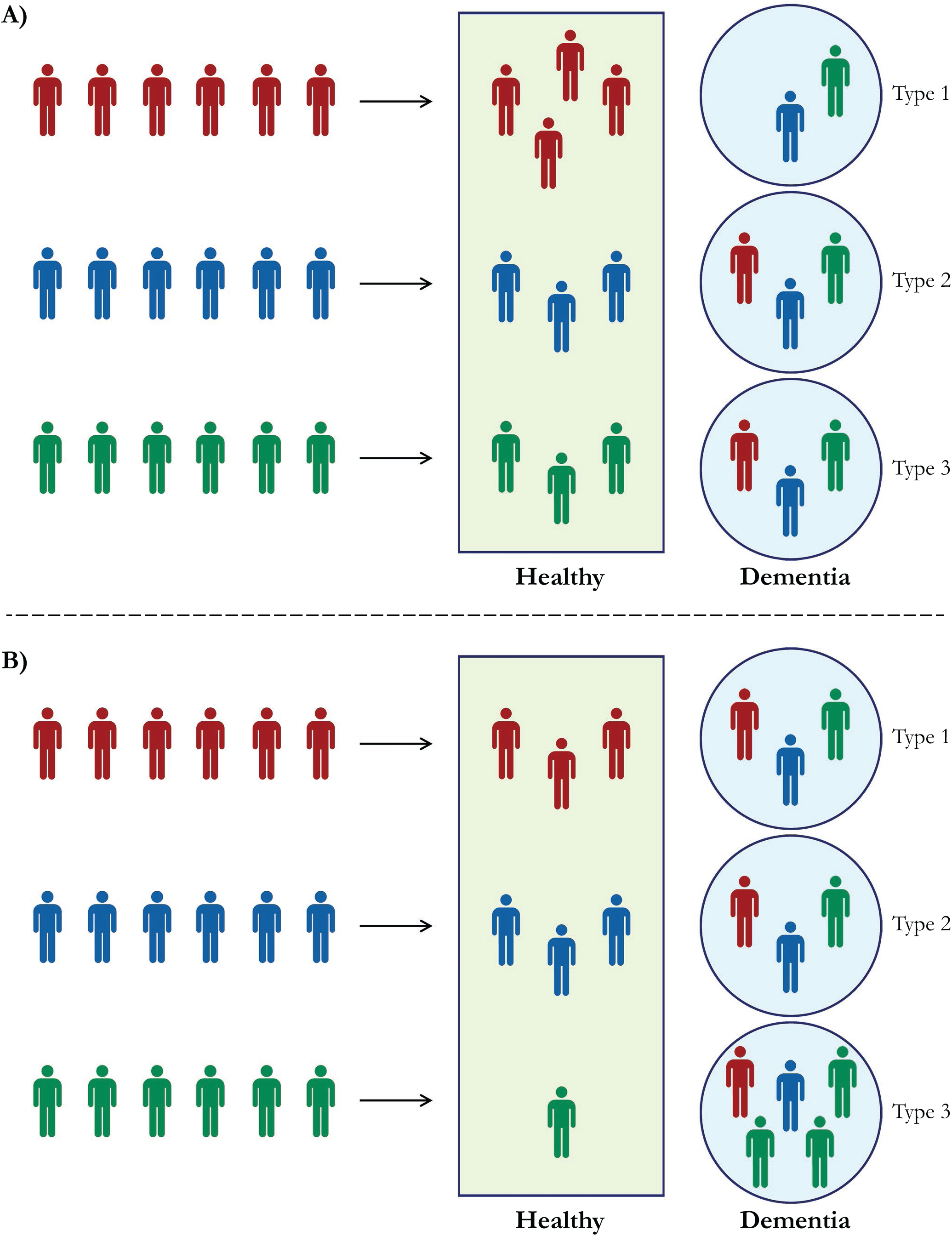
Schematic representation of protective and harmful effects and resulting cells in the contingency table. Each color (red, blue, green) represents a different occupational class. In scenario A, the red occupational class is relatively protected against dementia type 1, while the distribution of the remaining dementia types across occupations is equal. When the healthy control group is included in the analysis, this protective effect will be reflected in a higher proportion of individuals in red in the healthy group (4/6=67% versus 50% for blue/green) and a lower percentage of red persons in the type 1 dementia group (0% versus 1/6=17% for blue/green). All other proportions will be identical across occupational classes. However, when the healthy control group is not taken into account, the protective effect of the red occupation for dementia type 1 will create also an apparent “harmful” effect of the red occupation for dementia types 2 and 3 (i.e. 1/2=50%, compared to 1/3=33% for blue/green). In scenario B, the green occupational class shows a harmful effect on the development of dementia type 3, while no other differences between groups exist. Again, when the healthy control group is included, this only results in a lower percentage of healthy green individuals (1/6=17% versus 50% for red/blue) and a greater proportion of persons with green occupations in dementia type 3 (i.e. 50% versus 1/6=17% for red/blue). In the absence of a healthy control group, however, the green occupational class’ harmful effect for dementia type 3 leads to an apparent “protective” effect for dementia types 1 and 2 (1/5=20%, compared to 1/3=33% for red/blue).

